# Sport concussion in female athletes: a systematic review

**DOI:** 10.1101/2025.09.29.25336877

**Authors:** Ruby Westhead, Natasha Sigala

## Abstract

Concussions are very common injuries across many different sports and settings and present a significant public health concern in multiple countries. Previous studies have investigated concussion incidence, with a notable number reporting that female athletes have higher rates of sports-related concussions compared to their male counterparts. This study aims to look at risk factors linked to this increased susceptibility of female athletes, and how this affects them, rather than focusing on incidence.

To investigate this, we carried out a systematic review following the PRISMA guidelines. We searched the PubMed, Cochrane, APA PsychNet and Web of Science databases Inclusion criteria consisted of papers published in the last 10 years as of the 6th November 2023, studying athletes over the age of 18 who concussed themselves during sporting activity of any level. The exclusion criteria included narrative reviews, single case reports, abstracts, letters to the editor, and studies that included chronic traumatic brain injuries, spinal cord injuries or facial bone fractures.

After reviewing all the papers from the search (N= 1373), we identified 27 papers in total that met all inclusion criteria. We then extracted data from each paper relating to the biomechanics, neuropsychology, neurostructural substrates, physiology, and clinical recovery times. The main findings included women having significantly longer recovery times following concussions, increased symptom reporting, and different neurocognitive defects in testing post-concussion compared to men. There was also some evidence to suggest a difference in the neuroinflammatory process between men and women following concussive injury.

Highlighting these risk factors can hopefully lead to improved prevention, diagnosis, and treatment for concussions in women’s sports.

## Background

Sports-related concussions (SRC) are a type of mild traumatic brain injury (mTBI), induced by biomechanical forces that occur during a sporting event ^39^. This results in a range of signs and symptoms, one of which may be loss of consciousness ^39^. Due to this varied presentation, the acuteness of the injury, and the lack of reliable tests available, diagnosis can be difficult ^39^. In a nearly 4-year period spanning April 2017 to February 2021, the UK Government reported 7536 admissions to A&E with concussion, only 8.5% or roughly 640 of these were sport-related ^20^.

Concussions, particularly those obtained in sports, are a current focus in public health globally due to recent studies showing the long-term damage they can cause ^35, 37^. These include the risk of long-term neurodegenerative diseases increasing, including Alzheimer’s disease and chronic traumatic encephalopathy ^35^. The potential lifelong implication of the injury has led to legal action against Rugby Football Union and Welsh Rugby Union by former professional rugby players concerning lack of protection against long-term complications of concussive impacts^6^.

Diagnosis of concussion remains highly symptom-based^39^, making it one of the most complex sports-related injury diagnoses to make^39^. Despite these issues around concussion diagnosis, it remains common in sports ^27,52^. In Swedish elite soccer players, around 35% of them reported having an SRC at some point ^27^, with 10% having experienced one in the last year. This issue is exacerbated by athlete non-disclosure levels, with studies finding that athletes with a history of concussion have high rates of non-disclosure bias^2^. In the UK, most sporting bodies have guidelines outlining concussion management, including when to return to sport ^49^, but with diagnosis difficulty and participants avoiding disclosing symptoms, many still play on. Furthermore, these guidelines are rarely sex-specific^49^.

Many studies have shown an increased rate of concussions in female athletes compared to male athletes^13,15,55^. Van Pelt et al. reported that no matter the setting of injury, females were 2.02 times more at risk of sustaining a concussion compared to male athletes^55^. Cheng et al. found that in soccer and basketball, female concussion was significantly higher than male concussion incidence^13^. Not only are women more likely to sustain concussions, but they are also more likely to have more severe symptoms^32^ and a longer recovery period^5^.

Current research proposes several factors that may contribute to these sex differences in SRC. Some studies suggest the intrinsic differences between men and women - height, weight, head, and neck size - lead to greater SRC rates in women ^3,19 42^. Daneshvar et al. highlight that the difference between reporting behaviours in men and women explains a greater SRC incidence in men and women^19^, with males more likely to not disclose symptoms than females^31^. A link between the high symptom severity reported in female athletes and the normal hormonal changes caused by the menstrual cycle has also been suggested^11^.

Although many studies show females have an increased incidence of SRC, the focus of research in this field still is on male athletes and male-dominated sports. For example, D’Lauro et al. found that studies looking at female and male concussions focused on a population that was 80.1% male^18^. This lack of parity in female representation can lead to guidelines and management principles that may not be helpful to all athletes ^49^. The limited research into potential causes for the imbalance in rates of concussion leaves ambiguity for female athletes and their coaches on how best to protect them and help them recover from SRCs, with most research trying to generalise findings from male athletes for females.

This systematic review aims to further explore the reasons why female athletes are more susceptible to sports-related concussions than male athletes. With better knowledge of the predisposing factors to this imbalance, we aim to highlight areas where prevention, diagnosis and management can be improved for female athletes. Furthermore, by reviewing the relevant studies and looking at their limitations, we can emphasise where further research is needed in this field.

## Methods

This systematic review was carried out in accordance with PRISMA guidelines^45^ (Figure 1). The protocol was registered on the National Institute of Health Studies PROSPERO database and is available at: https://www.crd.york.ac.uk/PROSPERO/display_record.php?RecordID=473113

**Figure 1.**
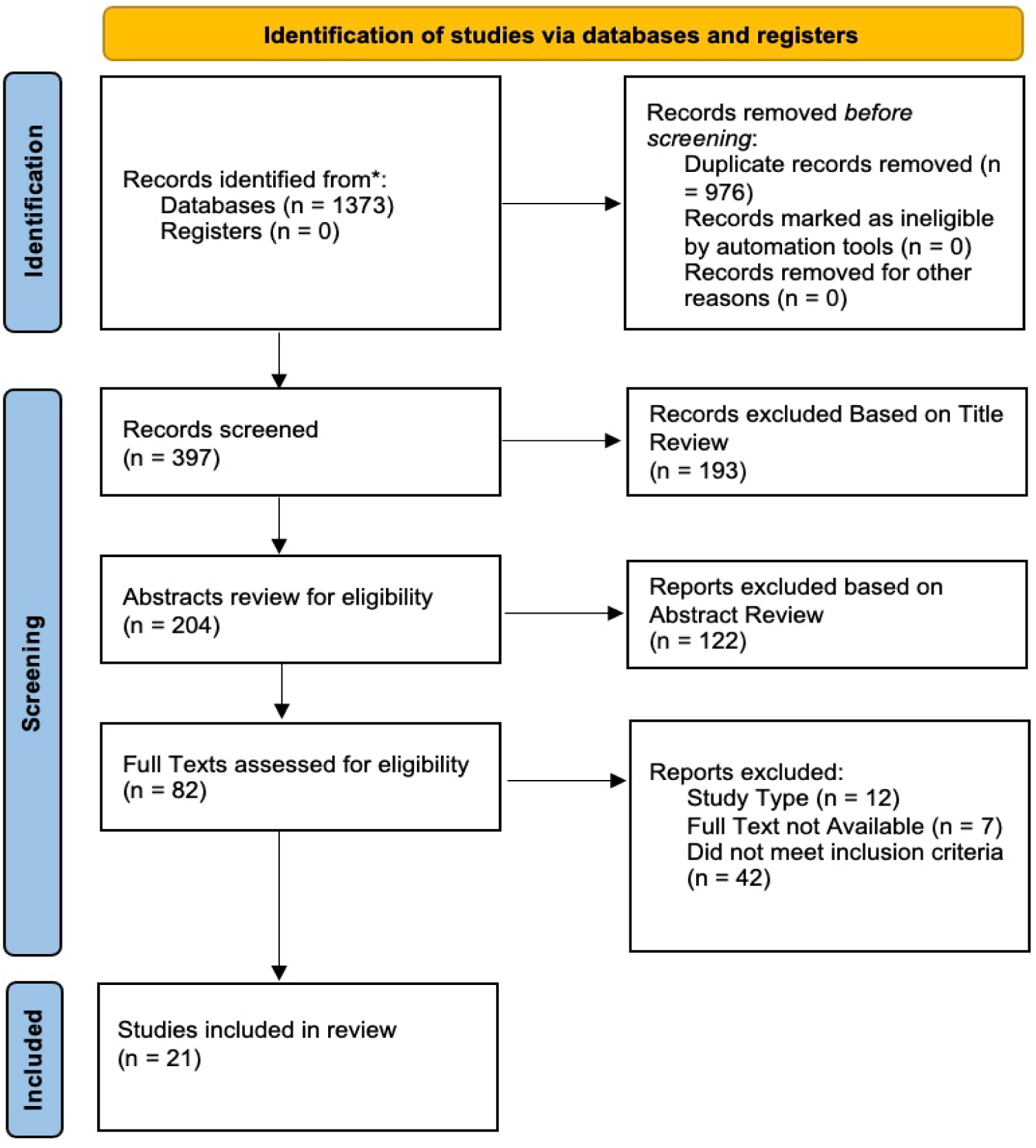
PRISMA flow diagram showing search strategy.

### Search Strategy

A search of PubMed, Cochrane, Web of Science and APA Psychnet databases was carried out on the 6th of November 2023. This search was done for each database using the following key terms: Concuss* AND Sport* AND Femal*. Each search was filtered to show articles published between 2013 and 2023, in English and with full text availability. These initial searches yielded a total of 1373 articles. From each database the articles were extracted into EndNote, where duplicates were deleted, leaving 397 papers for review.

### Inclusion and Exclusion Criteria

We used the following inclusion criteria: studies that focus on adult athletes who have sustained concussions in sports, published between 2013 and 2023 in English. Additionally, studies had to focus on concussions that were obtained during sporting activity and did not only find the incidence of sports-related concussions. The types of studies that could be included were cohort, case-control, and cross-sectional studies. Studies that included those with chronic TBIs, and those with facial fractures or spinal cord injuries were excluded. Also, narrative reviews, single case reports, abstracts and letters to editors were excluded from the systematic review.

Initially, the article titles were reviewed based on the above criteria, and then the abstracts. Finally, the methods and full texts of the remaining articles were reviewed. Figure 1 shows the full breakdown of the search process, at the end of which 21 articles remained in the review. Relevant information for these papers was extracted in an Excel spreadsheet (Table 1). Additionally, the following variables were extracted: (1) Clinical Recovery time, (2) Neurostructural substrates and physiology, (3) Biomechanics, (4) Neuropsychology and (5) Other, including results that could not be further grouped.

**Table 1.** Studies included in this systematic review. N: number of athletes / concussions, F: Female, M: Males.

| Lead Author (year) | Study Design | Sample | QualSyst Score |
| --- | --- | --- | --- |
| Bretzin (2021) <sup>9</sup> | Descriptive epidemiology study. | Concussed Collegiate Athletes<br>Men, N = 1209, Women, N = 765 | 0.83 |
| Churchill (2021) <sup>14</sup> | cohort study | University athletes. Concussed N = 61, Control N = 167 | 0.75 |
| Covassin (2016) <sup>17</sup> | Descriptive epidemiology study. | Collegiate Athletes in soccer, basketball, ice hockey, lacrosse, softball or baseball<br>Men, N = 779. Women, N = 903 | 0.67 |
| Di Battista (2016) <sup>22</sup> | Cross-sectional study | Concussed University athletes. N = 87, F = 27, M = 60 | 0.79 |
| Di Battista (2020) <sup>23</sup> | Cross-sectional study | Concussed university athletes. N = 40, M = 20, F = 20 | 0.75 |
| Esopenko (2020) <sup>25</sup> | Cohort Study | Collegiate athletes, N = 324<br>Concussed N = 56, M = 36, F = 20 | 0.67 |
| Heron (2023) <sup>29</sup> | Retrospective review | British Elite Cyclists<br>Concussed Athletes, N = 88, M = 54, F = 34 | 0.71 |
| Lumba-Brown (2020) <sup>36</sup> | Retrospective study of database | Concussed Athletes. N = 177, M = 103, F = 74 | 0.67 |
| McAllister (2023) <sup>38</sup> | Prospective multisite observational study | Concussed Collegiate Athletes and Military Cadets. N = 1751 | 0.83 |
| Mondello (2020) <sup>40</sup> | Prospective study | Collegiate athletes. N = 83, M = 41, F = 42 | 0.67 |
| Nolan (2023) <sup>43</sup> | Cross-sectional study | Collegiate athletes. N = 1104 | 0.79 |
| Oldham (2020) <sup>44</sup> | Prospective longitudinal | Collegiate Athletes. Concussed N = 50, Control, N = 25 | 0.79 |
| Schroeder (2022) <sup>48</sup> | Descriptive epidemiology study. | Water polo players, Concussions N = 83 | 0.67 |
| Shultz (2022) <sup>50</sup> | Prospective study | Amateur Australian Rules football players<br>N = 127, M = 82 F = 45. Concussed = 28 | 0.71 |
| Studenka (2019) <sup>54</sup> | Cross-sectional study | Collegiate Athletes<br>Hx of Concussion N = 73, M = 51, F = 22<br>Control N = 75, M = 49, F = 26 | 0.77 |
| Van Pelt (2021) <sup>56</sup> | Multi-site prospective cohort study | Ice Hockey Athletes<br>N = 332, M = 250, F = 82. Concussed N = 47 | 0.79 |
| Vedung (2020) <sup>57</sup> | Prospective cohort study | Elite Soccer Athletes<br>N = 959, M = 570, F = 389<br>Concussions, N = 36 | 0.77 |
| Walton (2020) <sup>58</sup> | Case control Study | Concussed Collegiate athletes. N = 20 | 0.83 |
| Walton (2020) <sup>59</sup> | Case control Study | University athletes. Concussed N = 28, control N = 28 | 0.79 |
| Weber (2020) <sup>60</sup> | Descriptive epidemiology study. | Female Collegiate Soccer. N = 381. Concussions, N = 25 | 0.67 |
| Wright (2022) <sup>62</sup> | Cohort study | Amateur Australian Football athletes<br>Concussed, N = 13 M = 7 F = 6. Control n = 16 M = 9 F = 7 | 0.67 |

### Assessment of risk of bias

Before results were synthesised, we reviewed each of the articles that matched all inclusion criteria on the quality of the study, using the Standard Quality Assessment Criteria for Evaluating Primary Research Papers from a Variety of Fields ^33^, also known as the Qualsyst Tool. This is a 14-point checklist, and we used an inclusion threshold of 0.55. No studies were excluded from our review based on their bias score (Table 1).

## Results

All papers and extracted information are presented in Table 1.

### Clinical Recovery Time

Our search identified 7 studies that investigated the difference of recovery durations between male and female athletes. Two of these focused on collegiate athletes of different sports ^9, 38,^ others focused on participants in specific sports, such as football ^17, 57, 60^, ice hockey ^17,56^ cycling ^29^, basketball ^17^, lacrosse^17^ and softball/baseball^17^. Bretzin *et al* studied a population of collegiate athletes in the USA who sustained concussions between the 2013-2014 and 2018-2019 academic years^9^. The study aimed to look at time taken for symptom resolution, return to academic life and return to play (RTP) - both limited and full return. RTP refers to resuming participation in sporting activities following an SRC. Each college’s athletic training team provided the data. The study found that on average women’s time for symptom resolution and return to academic life was significantly longer than their male counterparts^9^. It took 9 days for symptom resolution in women compared to 8 days in men, a difference that was statistically significant with a p-value of 0.03. With men taking 2 days less to return to academic life than women (7 days compared to 9 days respectively, p = <0.001 )^9^.

McAllister *et al.* looked at concussed collegiate athletes from both standard universities and military universities between 2014-2018. They found that when comparing concussed athletes with normal recovery times and slower recovery times, female athletes were more likely to be in the slow recovery group^38^.

When looking at concussions in female football players^17,57,60^, a wide range of results were found. Two studies^17, 57^ found that women had a significantly longer clinical recovery in some areas than their male counterparts. Covassin et al. reported that if the injury that caused the concussion happened in practice, females took longer to RTP than male athletes^17^. With the mean number of days out of play, 9.33 for women, and 6.14 for men being statistically significant (p = 0.006)^17^. Vedung et al found that female football players had a longer duration to RTP, which did not depend on where the injury was sustained (p = 0.02)^57^. Weber et al looked at female collegiate football players between the academic years 2004 - 2017^60^. They found that on average, concussions led to 3.96 missed games and 12.46 missed practices^60^.

Van Pelt et al look at collegiate ice hockey players using data from the CARE consortium, a multi-site prospective study looking into concussions in sports^56^. They reported that female ice hockey players took significantly longer to become asymptomatic compared to male ice hockey players^56^. On average, female ice hockey players were reported as being asymptomatic after 18.50 days, compared with males being asymptomatic after 6.85 days (p = 0.015)^56^. They also took longer to return to full participation in sports than their male counterparts (27.40 days vs 10.95 days respectively, p = 0.005)^56^.

One study looked at elite British cyclists between 2017 - 2022^29^. Over the course of this period, 88 concussions occurred, with female athletes being affected in 34 of these^29^. They found that even though for women the median RTP was 17.5 days, while for men it was 15.5 days, there was no significant difference^29^. Similarly, Covassin et al found no significant difference in clinical recovery times between the sexes in collegiate basketball, baseball/softball, ice hockey or lacrosse players1^7^.

### Neurostructural substrates and physiology

We identified 8 studies that investigated the effect of sports-related concussions on the structure,^14, 62^ neural substrates,^22, 23, 40, 50^ and physiology^58,59^ of the brain.

Churchill et al. and and Wright et al. used Magnetic Resonance Imaging (MRI) scanning to look at the structure of the brain post concussions^14,62^. Churchill et al. imaged their participants at acute injury time, RTP and 1 year following RTP^14^. They imaged both concussed athletes and a control group at the start of their competitive season. In terms of relative cerebral blood flow (CBF), they found that male athletes had a greater reduction in occipital-parietal CBF than female athletes^14^. Male athletes also had an increase in callosal mean diffusivity^14^. Female athletes had a decrease in fractional anisotropy (FA) in the corona radiata when compared to the female control group, with male concussed athletes on the other hand having raised FA compared to the male control group^14^. Wright et al, on the other hand, imaged concussed participants on day 13 following injury and compared them to a control group^62^. They found that straight sinus susceptibility values were decreased in concussed female athletes when compared to male concussed athletes^62^.

Four studies explored how the levels of different substrates were affected by SRC and if this showed a difference between the sexes^22,23,40,50^. One investigated biomarkers in those with a history of concussions and the second, the relationship between symptom burden and systemic inflammation between the sexes. In the first, they found that between the sexes different chemokines were raised in those with a history of previous concussions^22^. Female athletes’ levels of MCP-1 were higher in those with SRC history than females with no history of concussion^22^. For male athletes, MCP-4 concentrations were higher in the concussed group than in the healthy control group^22^. In the other study, Di Battista et al examined different cytokines and chemokine levels following SRC^23^. They found that male athletes’ symptom severity was linked to concentrations of IFN-gamma, whilst female athletes’ symptom severity was linked to MCP-4, and negatively correlated with IFN-gamma and TNF-alpha^23^. Additionally MCP-1, MIP-1beta and eotaxin levels were significantly lower in female athletes with SRC than male athletes with SRC^23^.

Mondello et al investigated T-tau levels following SRC^40^. They found that while in both sexes T-tau levels dropped, the lowest levels were found at different time points between the sexes following concussion^40^. Male athletes had the lowest level on day 3 post-SRC, whilst female athletes, following an initial rise, dropped on day 2, and then remained stably low^40^. Levels of T-tau in total were consistently higher post-SRC in female athletes compared to males^40^. Shultz et al focused on the levels of miRNAs following concussion^50^. MiR-21-3p and miR-223-3p levels were significantly higher in females than in male athletes (p = 0.0361 and p = 0.007 respectively)^50^.

The final two studies looked at how SRCs affected physiology and if there was a difference in this effect between male and female athletes^58,59^. Walton et al looked at the resting metabolic rate (RMR), percentage carbohydrate use (%CHO) and energy balance following SRC^58^. These variables were measured at 3-time points: acutely (≤ 72 hours), 7 days later, and after symptom resolution. They identified no significant differences in RMR/kg or %CHO between the sexes^58^. In male athletes, they reported that an increase in percentage carbohydrate use was positively linked with a longer time to becoming asymptomatic and RTP (p = 0.038 and p = 0.021 respectively)^58^. This was not the case with female athletes^58^.

Another study investigated how energy expenditure and energy balance are affected in the acute phase of SRC^59^. Concussed females had higher energy balance than female controls in the first 72 hours (p = <0.01), whilst there was no difference in the male athletes when compared to their matched control group^59^.

### Biomechanics

Esopenko et al focussed on the biomechanical differences between female and male athletes who suffered SRC^25^.

They examined the relationship between neck circumference and concussion rates^25^. Participants in this study were all collegiate athletes between 2013-2016^25^. Neck circumference was taken as they joined college and then they were followed up for any concussions they suffered during sports at college. They were also asked about their history of SRC before joining^25^. They found that males had larger neck circumference but there was no significant difference in circumference and SRC rates in college^25^. However, they found that female athletes who played sports with high risk for concussion had smaller proportional neck circumferences than those who played low-risk sports (p = 0.01) ^25^. This was not the case in male athletes^25^.

Proportional neck circumference was recorded to consider each athlete’s overall body mass, it was calculated by dividing the individual’s BMI by the neck circumference measurement ^25^.

### Neuropsychology

Three studies examined neurocognitive function following an SRC ^36, 44,54^. Lumba-Brown et al performed two cognitive tests. The first was the complete Sports Concussion Assessment Tool Version 5 (SCAT5) and the second was the Vestibular/Ocular-motor screening (VOMS) on concussed university athletes within 72 hours of injury^36^. SCAT5 is a concussion assessment tool that involves scoring post-concussion symptoms, memory, cognition, the Glasgow Coma Score, and balance ^36^. VOMS, on the other hand, involves measuring how post-concussion symptoms are induced whilst testing balance, vision and movement^36^, with patients rating symptom levels before and after each task. In this study, the tests performed in VOMS were related to smooth pursuit, horizontal and vertical saccades, convergence, horizontal and vertical Vestibulo Ocular Reflex, and visual motion sensitivity^36^. They found that female concussed athletes had significantly more atypical VOMS results in smooth pursuit (p = 0.045), convergence (p = 0.031) and visual motion sensitivity (p = 0.023) when compared to male SRC athletes^36^.

Studenka et al looked at non-linear motor performance on a seated visual-motor tracking task and how it is affected by concussions^54^. In this study, participants were asked to press a load cell with their finger at a force that matched the red line displayed on a monitor in front of them^54^. Their force was represented by a white line on the monitor^54^. Before starting the task, the participant’s maximal finger force was measured which allowed an estimate of the maximum voluntary contraction (MCV), 10% of this value was used to give the value for the red target wave line^54^. From this, the authors examined three different measurements: root-mean-square error score (RMSE), sample entropy (SampEn) and Average power (AvP) between 0 - 12 Hz ^54^. RMSE gave an overall metric of task performance, with a higher RMSE correlating to a poorer performance. This score represented the number of pixels on the monitor that the participants’ white line deviated from their target red line.

Sample entropy was used to demonstrate the regularity of the force output from each participant. A greater regularity in motor performance, or a lower SampEn score, shows how the participants had to have more attention to perform the task and so is linked to a general physiological impairment when carrying out the task. They discovered that females with a history of 2 or more concussions had a lower SampEn than females with no history of concussion (p = 0.001) and a history of 1 concussion (p = 0.026) ^54^.

The final measurement was Average Power. To allow for a measurement to be taken from the signal produced by the load cell, MATLAB was used. Using the Fourier transform, a formula that can reveal a signal’s frequency components, the square of the amplitude of the signal gives the average power at a given time series. The average power within three different bandwidths: 0 - 4 Hz, 4 - 8 Hz and 8 - 12 Hz was examined. Females with 2+ concussions had a lower average power in the 8 - 12 Hz range than females with 0 history (p = 0.043)^54^. Females with a history of 1 concussion had significantly lower AvP in the 4 - 8 Hz range than males in this category (p = 0.042)^54^. This trend continued for females with a history of 2 or more concussions when compared to males in this category (p = 0.031)^54^.

Finally, Oldham et al focused on tandem gait following concussion^44^. Participants, who were college athletes, performed either a single task (ST) or dual task (DT) at baseline and then 72 hours post-SRC^44^. Those in the control group had a baseline on a decided day and then a second test 72 hours later^44^. ST tandem gait testing involved asking the participants to walk in the heel-to-toe gait pattern along a 3m line before turning and walking back^44^. Anytime they failed to walk in the correct gait, or they did not follow the line this was considered a failed attempt, and they repeated the trial^44^. For DT, the same gait test was performed but the participants were asked to perform a cognitive task simultaneously^44^. This involved them doing either of the following tasks: spell 5 words backwards, subtract 6 or 7 from random numbers, or say the months of the year backwards^44^. Oldham et al found no significant difference between the sexes in either ST or DT (p = 0.99 and p = 0.53, respectively)^44^.

### Other

Finally, 2 papers provided useful information on the differences between male and female concussions but could not be further grouped into the above areas we focused on^43,48^.

Nolan et al investigated different symptom clusters that could be found in the clinical picture of concussion and how they were associated with specific risk factors^43^. This study was part of the CARE consortium and used data from 1104 collegiate athletes^43^. After identifying 4 symptom clusters: vestibular-cognitive, migrainous, cognitive function and affective, it was reported that females were associated with increased symptoms being reported in all 4 clusters when compared to male athletes (p = < 0.001)^43^.

The last study looked into mechanisms of injury in concussion in water polo players between 2016 and 2021^48^. Schroeder et al found that male athletes were 3.3 times more likely to hurt themselves in off-season practice than female water polo players^48^. Moreover, male athletes were 2.9 times more likely to sustain an SRC through contact with another player^48^. The most common mechanism of injury for female water polo players was through contact with a playing device^48^, whilst for men it was through contact with another player^48^.

## Discussion

This systematic review synthesised evidence on whether and how female athletes’ concussions differ compared to male athletes’ concussions. Differences reported include duration of clinical recovery, neural substrates, physiological marker levels post-SRC, and cognitive differences between male and female athletes leading to different risks of sustaining and recovering from concussion.

### Clinical Recovery Time

The reviewed studies reported several ways in which clinical recovery times differ post-sports-related concussion in females compared to male athletes. Some studies found that women took significantly longer to become asymptomatic^9,56^, and longer to return to play^5,17,38,57^. However, other studies found that this difference between sexes was only apparent in specific sports^29^, or where the injury occurred^17^. This lack of consistency on whether women take longer to recover compared to men has been documented in the literature before^41^, although female sex is still listed as a risk factor for prolonged recovery by the American Medical Society for Sport Medicine^28^. A study by Broshek et al., 2005, highlighted the need for individualised recovery plans considering the athletes’ sex instead of generalised set times spent out of play and academics^10^. Taken together, these findings suggest that guidelines for concussion management and time out of practice cannot be standardised for sex and sport.

This prolonged period during which females experience symptoms following a sports-related concussion could have wider effects on the health and quality of life of the athletes. A study found that the rates of female athletes who suffer from post-concussion symptoms for excessive periods following injury are greater than males^46^. Emanuelson et al., 2003, found that those who reported a greater number of post-concussion symptoms reported significantly lower scores on SF-36, a quality-of-life questionnaire^24^. In this study, concussed individuals carried out the SF-36, alongside a post-concussion symptoms checklist, at 3 months and 1 year post injury^24^. Furthermore, this review found evidence that women spend a longer period out of academics^9^. With a lack of research in this area, it is not clear if there are long-term complications and so this is an area where further research is vital.

### Neurostructural substrates and physiology

Several papers examined concussions’ long-lasting effects on the brain, including its structure with imaging, and different physiological biomarkers of tissue inflammation. Two papers used MRI scanning in the acute phase of SRC, they showed differences in how the brain is affected between males and females. Churchill et al looked into how different areas of the brain are affected acutely following concussion and in the long term^14^. In this study, there was no sex difference in the clinical presentation of each participant’s concussion^14^. One main finding found that male athletes had a greater relative reduction in CBF in the occipital-parietal region of the brain^14^. This supported a previous studies findings^26^. Studies have found that this difference in how CBF is affected may be because the female sex hormone oestrogen is neuroprotective, allowing the brain to better maintain cerebral autoregulation^47^.

Overall, this helps maintain CBF levels and so could explain why Churchill et al, found that male athletes had a greater reduction^14^. However, more research is needed into how oestrogen is neuroprotective in concussions and the long-term effects of this protection in recovery. Future studies should also explore whether this is affected by the menstrual cycle and use of hormonal contraceptives. Potentially seeing if there are benefits in using oestrogen in males at high risk of concussion.

Furthermore, the same study found that females had decreased levels of FA in the corona radiata^14^. This could demonstrate how mTBIs can lead to white matter injury in the brain following neuroinflammatory glial activation^53^ and a suggestion of greater diffuse injury following a concussive impact in females^14^. It has also been shown that decreased FA is linked with worse long-term outcomes when it comes to recovery^34^. Finally, the differences in mean diffusivity could imply the difference in the biomechanics of concussive injury between the sexes. This is an area where further research is needed looking at how the brain is affected by different mechanisms of injury.

Additionally, this review found evidence about how physiological substrates differ in female and male concussed individuals. Though all studies found statistically different levels in many different substrates, there was no indication of how this affects the athletes’ clinical presentation or long-term recovery from concussions.

Proteins such as MCP-1 and MCP-4 were shown to be in different concentrations between the sexes^23^. These are two important chemokines in facilitating peripheral immune cell migration to the CNS following injury^23^. This finding shows that following concussion a neuroinflammatory response is due to tissue damage. More research is needed into whether these chemokine levels are due to them working to repair the tissue damage or if they are showing continued damage to the brain long-term. This study agrees with current research into the sex differences in the inflammatory response due to the sex hormones mediating production differently^7^. This sex difference in the neuroimmune response has been highlighted in other studies, Villa et al., 2018, found that microglia gene expression is based on an inflammatory phenotype in male mice compared to a neuroprotective phenotype in female mice^21^. However continued research is needed to explore the sex differences of the neuroinflammatory response, and how these differences can be used to better manage concussions in athletes. Due to no true comparison between inflammatory markers and symptom severity or clinical outcome of concussions, it is hard for this review to draw firm conclusions on how this could better manage female athletes affected by concussions.

Furthermore, one study reported that concussed females had a higher energy balance when compared to controls^59^. This difference between male athletes - where this is not applicable - again highlights the emphasis needed on sex-specific individualised recovery guidelines instead of broad programmes.

### Biomechanics

We identified one paper that discussed the biomechanics of concussion and how this could differ between the sexes. This found that, when controlling for sex, neck circumference had no association with SRC rates^25^. This study agrees with other current research that suggests neck strength is protective of SRC^51^, rather than the size of the neck itself. Studies found that females could be at a greater risk of concussion due to them on average having less overall neck strength when compared to their male counterparts^4^. However, this review found no data of significance to explain the difference in biomechanics of concussions between the sexes and so for this reason cannot comment.

### Neuropsychology

Our systematic review showed how important it is to administer neuropsychological tests to concussed people to ensure that the initial concussion is fully investigated, recovery is optimised, and the focus is not only on the initial symptoms. It also highlighted how important it is that male and female athletes are treated differently when concussed, as studies showed how the sexes can have different lasting neurocognition effects^36, 54^. Studenka et al, demonstrated the importance of using visual motor tracking tasks in female athletes to monitor for long-term visual-motor impairment following concussion^54^. This will be very important in aiding recovery pathways for female athletes and better protecting them from complications. This was due to the finding that female athletes may be more likely to have suffered longer-lasting changes in their visual-motor performance, shown by concussed females having a lower SampEn than females with 0 concussion history^54^. Further research into the mechanisms behind this pathological change is needed, however, to work out how recovery guidelines should change.

Average Power for females being lower in the 4 - 8 Hz frequency domain than males, show a higher impairment in the feedforward or predictive control motor control systems^21^. This once again shows proof of the long-term consequences in the visual-motor control systems suffered by female athletes. Overall, the effects of concussion on neurocognition in the long term is an important area of further research.

These findings agree with current knowledge into how visual-motor-tracking tasks change in females post concussions^30^ and suggest how female athletes may be more at risk of long-term changes to this^36^. Continued research into the pathophysiology of how concussions affect VOMS differently between the sexes is important to allow a move to a more personalised assessment and care in concussions. This care should consider including symptoms review, and neurocognitive and postural control assessments^39^ at the diagnosis stage that should be analysed at a sex-specific level.

### Other

Previous studies have discovered that concussions suffered by female athletes have a greater symptom burden than concussions suffered by male athletes^1,12^. Our review found one paper that matched this^43^, with female athletes being a risk factor for increased symptoms in all 4 clusters reported by Nolan et al. However, some research suggests that this could be caused by female athletes’ willingness to report symptoms more than male athletes^16, 31^. Due to only one study being found that spoke directly of symptoms, it is difficult to draw firm conclusions about it. However, with many studies suggesting it, it is important that in future literature this is further studied. This could be a significant area of management that should be changed for female athletes.

Schroeder et al focused on how the mechanism differs between concussive head injuries in the different sexes^48^. Williams et al., 2021, though not looking at concussions did look at head impacts in general in Rugby union, found substantial differences in how male and female players hit their head with uncontrolled whiplash occurring in >50% of female impacts and <0.5% of males^61^. Similar to Schroeder et al, other studies have found that females are more likely to suffer concussions from non-player contact compared to male athletes who suffer concussive impacts from contact with another player^8^. Nevertheless, there is a gap in research indicating how this could be explained and so further research is required, and how this information could be used to better protect women in sport.

### Limitations

There are several limitations to this systematic review.

Firstly, many of the studies we reviewed had sample sizes too small for their findings to be generalised to the whole population they were discussing (i.e. all female athletes in that sport(s). This limited the quality of many of the studies and could have affected the validity of the results they produced. This limitation to generalise any reported differences was made worse by the number of studies that had more male participants than female participants when making comparisons between them.

Secondly, due to the complex diagnostic features of concussion and the varied clinical presentations, data was hard to validate. Furthermore, outcome measures were variable across studies, so comparisons were done on a more general level of cognitive functions, instead of on specific tests.

Thirdly, when studies were focussing on clinical recovery time, they employed self-reported symptom levels to assess if they were symptomatic. Due to differences in how people self-report, these data may not be as reliable. Future studies could look at more standardised and accurate ways to check for recovery time using other tests rather than symptom questionnaires.

Many of the studies produced results based on American college athletes, with several studies being under the CARE consortium and so could have involved the same population. This makes it hard to say if the results are generalisable to the wider athletic population.

Finally, many studies relied on self-reported concussions. As discussed, it is known that men are more likely to not disclose concussive head injuries compared to females^31^. It could also mean data provided might not be the result of concussion but of other injuries. Due to this, future studies should ideally focus on cohorts of medically confirmed concussions.

## Conclusion

Our systematic review demonstrated that female athletes have longer recovery periods, increased symptom numbers and a worse neurocognition impact post-concussion than male athletes. It also suggested a difference in the neuroinflammatory response in females compared to males and varying changes to the brain structure that could be linked to longer clinical recovery. Further research is needed into SRC in female athletes to better protect, diagnose and treat these injuries.

## Data Availability

All data produced in the present study are available upon reasonable request to the authors

